# Frequent post-treatment monitoring of colorectal cancer using individualized ctDNA validated by multi-regional molecular profiling

**DOI:** 10.1101/2020.06.10.20126367

**Authors:** Mizunori Yaegashi, Takeshi Iwaya, Noriyuki Sasaki, Masashi Fujita, Zhenlin Ju, Doris Siwak, Tsuyoshi Hachiya, Kei Sato, Fumitaka Endo, Toshimoto Kimura, Koki Otsuka, Ryo Sugimoto, Tamotsu Sugai, Lance Liotta, Yiling Lu, Gordon B. Mills, Hidewaki Nakagawa, Satoshi S. Nishizuka

## Abstract

**Purpose:** Circulating tumor DNA (ctDNA) analysis has been proposed as an approach for prediction of post-treatment patient outcomes. However, whether a single platform will provide optimal information in all patients or alternatively a patient-specific monitoring approach based on assessment of mutations in the primary tumor from that patient remains an urgent question. Experimental Design: We conducted multiregional sequencing of 42 specimens of 14 colorectal tumors (Stage III and more) from 12 patients, including two double cancer cases, to identify the full spectrum of mutational heterogeneity and identify aberrations that could be used to develop personalized ctDNA assays.

**Results:** “Founder” mutations that occur in all regions of the sample were identified in 12/14 (85.7%) tumors. Subsequent phylogenetic analysis of each tumor showed that 12/14 tumors (85.7%) carried at least one “truncal” mutation. Most founder and truncal mutations exhibited higher variant allele frequency (VAF) than “non-founder” and “branch” mutations. In addition, both founder and truncal mutations were more likely to be detected as ctDNA than non-founder and branch mutations. Synchronized ctDNA dynamics of multiple mutations suggested those mutations from the same clonal origin. For 10/12 patients (83.3%) with nearly 1,000 days of post-operative observation, the validity of frequent personalized ctDNA monitoring was confirmed in terms of early relapse prediction, treatment efficacy, and non-relapse corroboration.

**Conclusions:** Personalized ctDNA monitoring based on aberrations with a high VAF in the primary tumor site should be explored in larger prospective clinical trials to determine the full clinical validity.

**Translational relevance:** Circulating tumor DNA (ctDNA) has been reported to be a new class of tumor-specific personalized biomarkers, but the selection criteria of index gene mutations from heterogeneous tumors as well as the achievement of sufficient sensitivity remain a challenge. Among mutations detected by multiregional sequencing, we monitored mutations with high variant allele frequencies (VAFs) from advanced colorectal cancers. Clinical validity of longitudinal ctDNA monitoring using highly-sensitive digital PCR was evaluated in terms of: (a) early relapse prediction; (b) treatment efficacy evaluation; and (c) no relapse corroboration. We found that ctDNA from high VAF mutations of a tumor are likely to be founder/truncal mutations. Based on rigorous longitudinal monitoring, our results suggest that sensitivity required the VAF to be 0.01-0.1%. The ctDNA from high VAF mutations strongly reflects tumor burden in a timely manner, thereby establishing clinical validity as a new class of tumor-specific personalized biomarkers.

## Introduction

Colorectal cancer (CRC) is the third most common cancer diagnosed worldwide with approximately 1,800,000 new cases and approximately 881,000 deaths in 2018 (1). Post-treatment relapse with distant metastasis (mCRC) represents the major cause of CRC-related deaths. The outcome for mCRC has improved dramatically over the last 20 years, particularly in terms of duration of survival, in part due to therapeutic improvements including less-invasive surgery and ablative techniques combined with emerging chemo and molecular systemic therapies (2). However, the best surveillance approach to determine the need for post-treatment interventions has not yet been established. Although recent clinical trials demonstrated that frequent surveillance using computerized tomography (CT) and/or serum carcinoembryonic antigen (CEA) could increase the likelihood of successful recurrent site resection with curative intent, such surveillance had very little overall impact on survival (3,4). These results suggest that earlier post-treatment interventions will be required to reduce CRC-related deaths.

Circulating tumor DNA (ctDNA) represents an emerging tumor marker (5). The potential utility of ctDNA is based on the principle that somatic mutations are derived exclusively from cancer cells and thus may facilitate earlier detection of post-therapy recurrence compared to conventional serum tumor markers and imaging approaches (6). In addition to recurrence monitoring, ctDNA can be used to select patients for molecular targeting drugs, particularly when biopsy samples cannot be obtained (7). A major challenge to the utility of ctDNA as an individualized tumor marker is the need to account for mutational tumor heterogeneity both between patients and within patients as a consequence of tumor evolution and treatment (8-11).

Although DNA fragments are released from heterogeneous tumor cells, whether mutational heterogeneity across a tumor is fully reflected in ctDNA remains unclear. In the present study, we first assessed mutational heterogeneity of tumor samples using multiregional sequencing followed by phylogenetic simulation analysis (12).

Another challenge of implementing ctDNA monitoring in daily practice is the identification of a sensitive and practical detection system for frequent monitoring of the very low variant allele frequencies (VAFs) present in blood. The detection limit of VAFs by next generation sequencing (NGS) is generally >1% (6). Indeed, the majority of ctDNA VAF is <1%, even in recurrent cases (13). In the present study, we implemented a highly specific and personalized digital PCR (dPCR) primer/probe approach using Hypercool Primer & Probe (HPP) technology to robustly detect mutations identified in resected tumor specimens from that patient (14,15). In our system, dPCR with HPP technology has a 0.01% VAF detection limit as well as >95% successful reaction rate The high dPCR success rate facilitates a cost effective and frequent patient-specific tumor burden monitoring approach in daily practice that can facilitate: (a) early relapse prediction; (b) treatment efficacy evaluation; and (c) non-relapse corroboration. Here we present the evaluation of genomic characteristics of post-treatment Stage III/IV CRC along with results for frequent ctDNA monitoring conducted over approximately 1,000 follow-up days.

## Materials and Methods

### Study design

The subjects enrolled in the present study were analyzed as part of an observational study. All enrolled patients had intention-to-treat for pathological Stage III or more advanced CRC and were treated at the Department of Surgery, Iwate Medical University School of Medicine between March, 2016 and December, 2016 (HGH27-29). In principle, all patients received treatment for advanced CRC according to Japanese Society for Cancer of the Colon and Rectum guidelines (17). No specific intervention and specifically-scheduled clinical examinations for this particular study were allowed. Pre- and post-treatment peripheral blood samples were obtained in addition to routine clinical laboratory examinations. All samples were acquired after obtaining written informed consent and approval by the Institutional Review Board of Iwate Medical University School of Medicine according to the Helsinki Declaration.

### Sampling and preservation

All primary tumor tissues were acquired postoperatively. Based on a macroscopic diagnosis of resected tumors, samples were taken from three regions in the tumor that were spaced at least five millimeters apart. Each sample was divided into two portions from which DNA was extracted and cell lysates were prepared before storage at −80 °C. The first whole blood sample was collected into a BD Vacutainer CPT blood collection tube (Becton, Dickinson and Company, East Rutherford, NJ). The whole blood sample was separated into plasma and peripheral blood mononuclear cells (PBMCs) that were used to analyze germline DNA. Within 2 hours of collection, both plasma and PBMC phases were further centrifuged separately at 1,800 *g* at room temperature to remove other components. The follow-up blood samples were acquired “frequently” (e.g., every 1-3 months) such that collection was carried out simultaneously with routine clinical-pathway laboratory examinations in 2 x 10 ml volumes in two Cell-Free DNA Streck BCT blood collection tubes (Streck, Omaha, NE) at room temperature. Within five days of blood withdrawal into a BCT tube stored at room temperature, the blood sample was centrifuged at 1,800 *g* for 20 min at room temperature to separate plasma and red blood cells. The plasma phase was transferred to another tube that was centrifuged at 1,800 *g* for 20 min at room temperature to remove cellular debris. The isolated plasma phase was stored at −80 °C.

### DNA extraction

The genomic DNA was extracted using a QIAamp DNA Mini Kit for tumor tissue and PBMCs; and a QIAamp Circulating Nucleic Acid Kit for plasma (Qiagen, Germany). The extracted DNA in solution was transferred to a 0.5 ml tube and stored at −30 °C until analysis. The quantity of extracted DNA was measured using a Qubit12.0 dsDNA high sensitivity assay kit (Life Technologies, Carlsbad, CA).

### Panel sequence

For each patient, genomic DNA was extracted from three regions of the primary tumor and from PBMCs. NGS libraries were prepared using ClearSeq Comprehensive Cancer Kits according to the manufacturer’s instructions (Agilent Technologies, Inc., USA). The ClearSeq Comprehensive Cancer Panel targets 151 disease-associated genes and was analyzed using an Illumina Hiseq 2000 (enrichment system) (Supplementary Table S1, Illumina, Inc., San Diego, CA). Qualification of variant calling was performed by adaptor-trimming of reads using Cutadapt (http://code.google.com/p/cutadapt/) and mapping to GRCh37 using Burrows-Wheeler Aligner (18). PCR duplicates were removed using Picard (https://broadinstitute.github.io/picard/). Low-quality reads were filtered based on mapping quality, number of mismatches and INDELs. Improper reads were filtered based on discordance among chromosomes, as well as direction and distance of paired-end reads. SNVs and INDELs were called using VarScan2 (19) with a minimum read depth of 20, a minimum variant allele frequency of 5%, minimum supporting reads of four, and a p-value threshold of 0.05. The variants were annotated using Ensembl VEP. Copy number (CN) analysis was performed using VarScan2 and DNAcopy (20).

### Copy number variation

Copy number variation (CNV) was calculated using ONCOCNV obtained via GitHub (21), with BAM files as input. Read counts in tumor BAM files were normalized, corrected for colon cancer-content, and the CNV was detected by comparison with the baseline copy number. The baseline copy number (CN) was defined based on PBMC BAM files and was subsequently used for CNV calculation for all multiregional samples. CN segmentation was performed using the DNAcopy package of R/Bioconductor. ONCOCNV was run on the SHIROKANE supercomputer at the University of Tokyo Institute of Medical Science.

### Phylogenetic tree

Phylogenetic trees were constructed using a modified version of Canopy (version 1.3.0), an open source R package (https://cran.r-project.org/web/packages/Canopy/) (12). A list of somatic SNV/INDELs was used as input. Based on preliminary simulations of evolutionary trees, SNV data were prioritized for the simulation in the present study in which CNV data were limited to those from the panel sequencing. Clustering of SNV/INDELs was performed during preprocessing to accelerate simulation convergence. Markov chain Monte Carlo simulations were run 10 times, and the maximum simulation length was 100,000 steps. Convergence of simulations was confirmed by visually inspecting the time course of log likelihood and acceptance rate in all cases. Data from the burn-in phase were discarded.

### RPPA

Tissue samples were serially diluted two-fold (undiluted, 1:2, 1:4, 1:8, and 1:16) and arrayed on nitrocellulose-coated slides to produce sample lysate spots. Signals from the sample spots were then developed via an immunochemical reaction and tyramide-based signal amplification before visualization with a GenPoint DAB colorimetric reaction (Agilent Technologies, Santa Clara, CA). The developed slides were scanned on a Huron TissueScope scanner to produce 16-bit TIFF images. Sample spots in the TIFF images were identified and their densities were quantified using an Array-Pro Analyzer (Meyer Instruments, Houston, TX). Relative protein levels for each sample were determined by interpolating each dilution curve produced from the densities of the 5-dilution sample spots using a “standard curve” (SuperCurve) for each slide (per antibody) (22). A SuperCurve was constructed using a script written in R (The R Foundation for Statistical Computing, Vienna, Austria). All data for relative protein levels were normalized for protein loading and transformed to linear values, designated as “Normalized Linear”, which were transformed to log_2_ values before being median-centered for hierarchical clustering analyses. The heatmaps were developed at: Department of Bioinformatics and Computational Biology, University of Texas MD Anderson Cancer Center; In Silico Solutions (Falls Church, VA); Santeon (Reston, VA); and SRA International (Arlington, VA).

### dPCR

Mutations in *KRAS* and *BRAF* and *PIK3CA* were analyzed using a commercially-available primer/probe kit for dPCR (Thermo Fisher Scientific). Primer/probe sets for SNVs were designed and synthesized using Hypercool^™^ Primer & Probe technology (Nihon Gene Research Laboratories, Inc., Sendai, Japan) wherein forward and reverse primers and a hydrolysis probe were designed that would produce an amplicon of ∼70 bp. Several adenine or cytosine bases in these primers/probes were replaced with “2-amino-dA(2aA)” and “5-Methyl-dC(5mC)”, respectively, to ensure a high Tm value despite the short amplicon length. Primer/probe sets originally designed using Hypercool^™^ Primer & Probe technology were validated for use with dPCR and DNA from primary tumors (Nihon Gene Research Laboratories, Sendai, Japan). The QuantStudio 3D Digital PCR System (Thermo Fisher Scientific) was used for PCR and counting of absolute mutation fragments. The maximum input of plasma cfDNA ranged from 0.2-8.5 µl per dPCR assay. VAF was calculated when at least two mutant-type signal dots (FAM) at one time point or more than one mutant-type signal dot at consecutive time points existed in the presence of wild-type signal dots (VIC) using the formula:

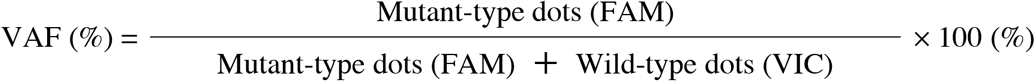

### Statistical analysis

Both parametric and nonparametric tests for group comparison were performed, and included Student’s t-test, Wilcoxon signed-rank test, Mann–Whitney U test for continuous variables; chi-square test, and Fisher’s exact test for categorical variables; and Pearson’s or Spearman’s correlation coefficient for two variables. All statistical analyses were performed using JMP software version 14.0 (SAS Institute, Inc., Cary, NC, USA). A probability (*p*) value of <0.05 was considered statistically significant.

## RESULTS

### Patient characteristics

In our prospective observational study, 44 clinical Stage II-IV CRC patients with disease that was considered to be resectable at the time of diagnosis were registered between March 2016 and December 2016. Two patients were excluded due to unresectable lesions and pathological Stage I disease. The other 42 patients that were confirmed to have pathological Stage II-IV CRC underwent primary tumor resection. For the present study, 12 patients were enrolled based on the following criteria: (a) availability of three samples collected from a primary tumor that was at least pathological Stage III; and (b) confirmation of tumor cellularity >40% in all specimens (Supplementary Fig. S1, Supplementary Table S2). The final number of tumor regions sequenced was 42, which were taken from 14 tumors obtained from 12 patients. Among these patients, two patients had two separate colorectal tumors.

### Multi-regional sequence of primary tumors by NGS

Sequence analysis was performed using the ClearSeq Comprehensive Cancer panel that targets 151 disease-associated genes (Supplementary Table S1). A total of 157 mutations from 84 genes were identified in the 14 primary tumors from 12 patients, including two double cancer cases (Fig.1). The average number of mutations per tumor was 12.1 [range: 3-55], whereas the average in a single region was 7.2 [range: 2-43]. For this study, a founder mutation was defined as a mutation that is present in three regions of the tumor; 12/14 (85.7%) tumors had founder mutations. Patients CC16011 and CC16023 had no founder mutations but did have 40 and 55 non-founder mutations, respectively. In the 42 samples examined, 156 founder mutations (45 unique mutations) and 147 non-founder mutations (113 unique mutations) were identified. The average number of founder and non-founder mutations per tumor was 3.7 [range: 0-9] and 8.4 [range: 0-55], respectively. The median VAF (%) of founder mutations was significantly higher than that of non-founder mutations (30.0 [IQR; interquartile range: 22.9-41.4] vs. 22.4 [IQR:12.1-38.8]; p<0.0001; Supplementary Table S3).

### Phylogenetic trees for primary tumors

Phylogenetic trees were generated to simulate cancer clonal composition and chronological evolution based on a multi-regional mutation profile (12). A simulation for CC16001 yielded multiple clonal compositions in an individual region and also showed the relative timing of introduction of new mutations (Fig. 2A). To understand chronological evolution, here, “truncal” mutation was defined as the first mutation that occurred prior to divergence of branches in the phylogeny tree. The median VAF (%) of truncal mutations (71.5[IQR: 55.7-80.5]) of CC16001 was significantly higher than that for branch mutations (26.7[IQR:20.1-37.1]) (p<0.0001). The average number of truncal mutations per tumor was 1.4 [range 0-3], whereas the average for “branch” mutations was 72.6 [range 13-464]. As predicted, in most tumors, the median VAF (%) of truncal mutations (48.6 [IQR: 37.1 – 60.9]) was significantly higher than that for branch mutations (21.2 [IQR:9.4 – 30.7]) (p<0.0001, Supplementary Table S4). Analyses for all tumor samples are shown in Fig. 2A and Supplementary Fig. S2.

**Figure 1.**
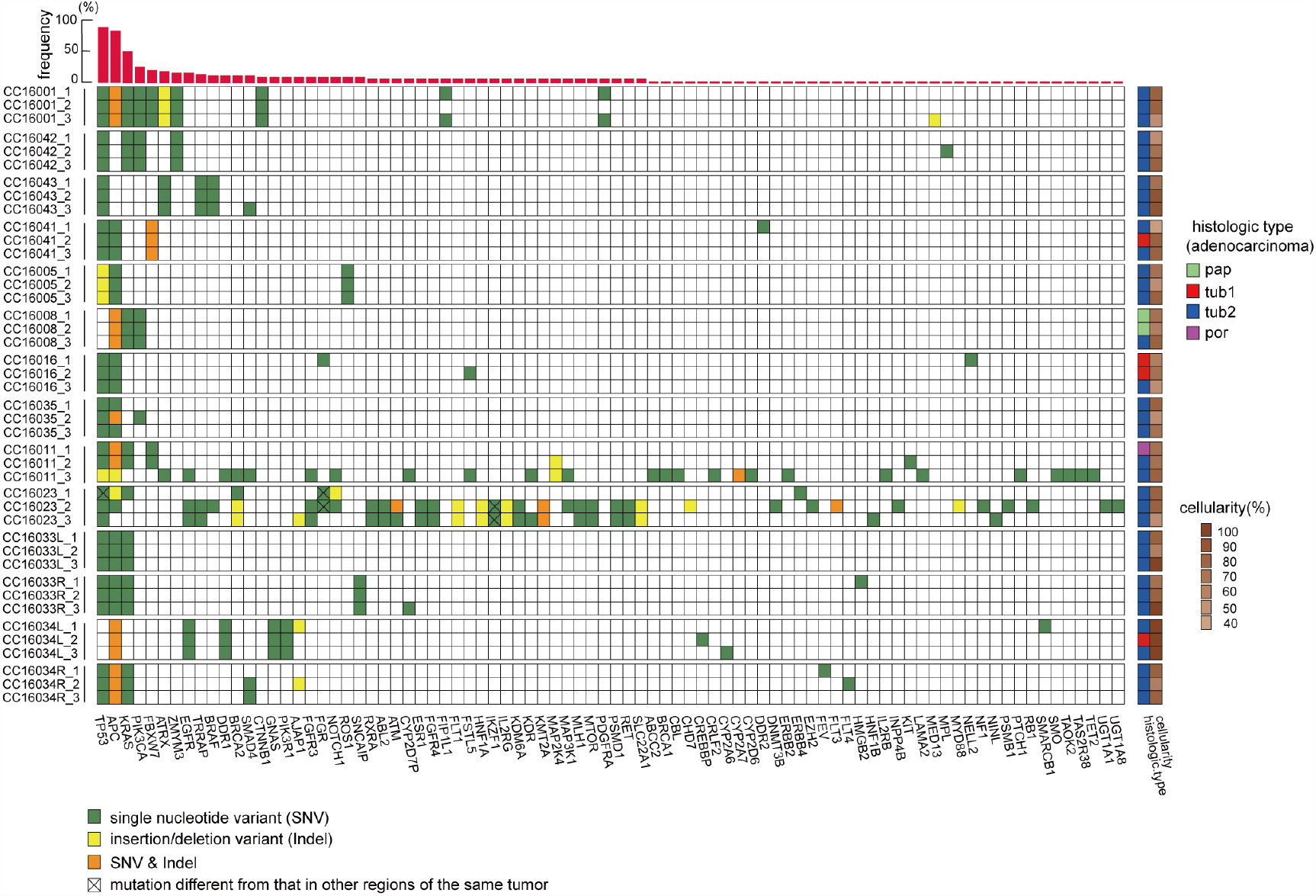
Multi-regional sequencing of primary tumors. The horizontal axis shows genes that were mutated in at least one sample and the vertical axis lists case number and sample region. Green and yellow squares indicate a single nucleotide variant (SNV) and insertion/deletion variant (INDEL), respectively. Orange squares indicate the presence of both SNV and INDEL. The same color square for a gene in identical tumors indicates the presence of the same genetic mutation; however, the x-mark in the same color squares mean that these mutational locations are different from each other. These mutations show non-synonymous variants. The red bar graph at the top indicates the frequencies of mutations in 42 sample regions. Light green, red, blue, and magenta squares in the right hand column indicate papillary, well-differentiated, moderately-differentiated, and poorly-differentiated adenocarcinomas, respectively, along with the cellularity of the corresponding regions.

**Figure 2.**
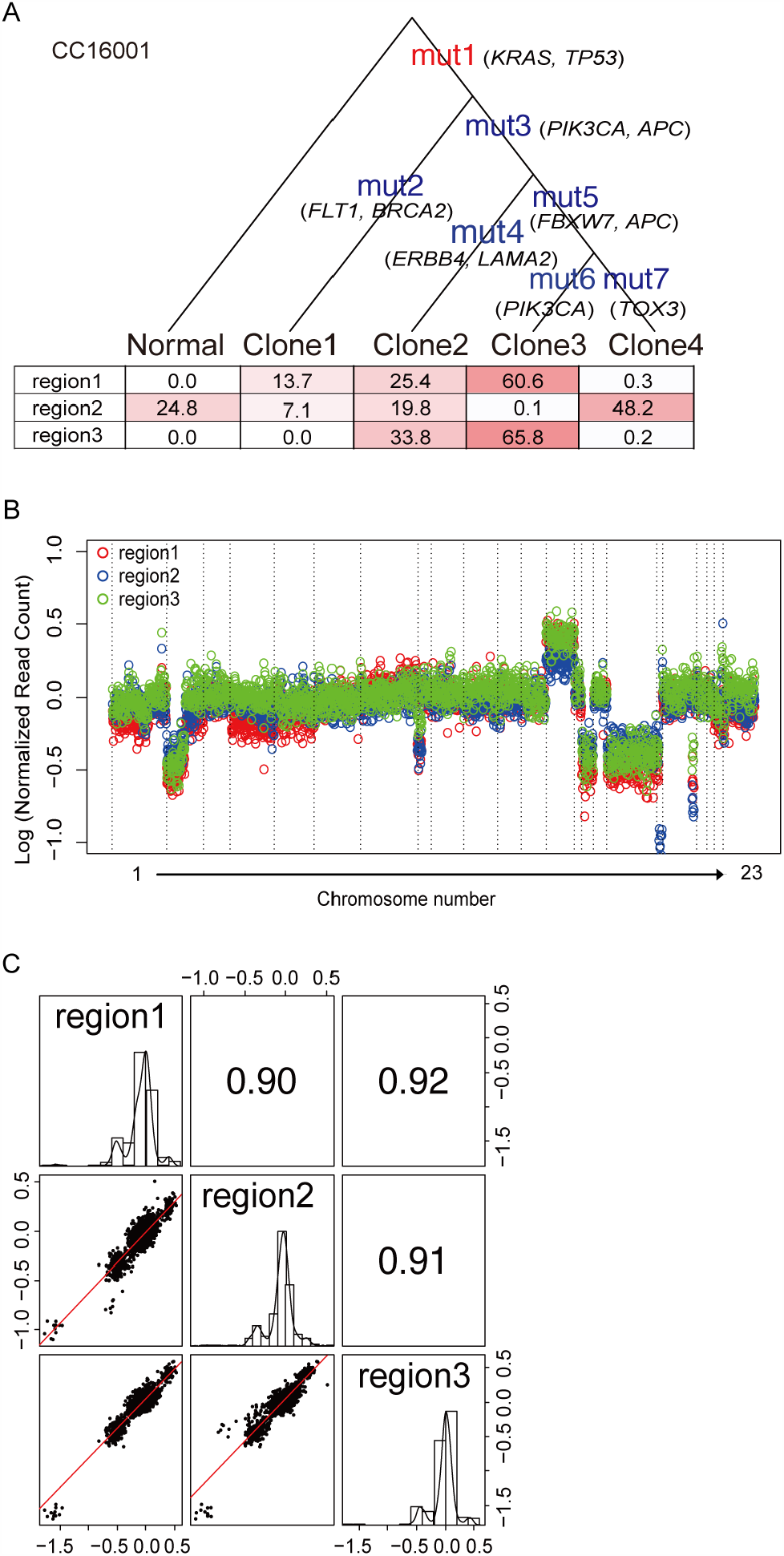
Intra-tumor genetic heterogeneity represented by phylogenetic tree and CNV. (A) Letters in red and blue indicate truncal mutations and branch mutations, respectively. The tables at the bottom of the phylogenic tree show simulated proportions (%) of each clone for three sample regions of the primary tumor. The set of mutations per sample is provided in Supplementary Data file S2. (B) Color dot representation of copy number across chromosomes in three regions of a tumor taken from CC16001. (C) Pearson’s correlation coefficients for all possible CNV combinations between three sample regions. Scatter plots are of two CNs of a given pair, whereas histograms show the frequency of CN distribution of a region.

### Founder and truncal mutations

The founder mutation is defined in a binary manner (i.e., presence or absence), whereas the truncal mutation is a result of statistical simulation in a quantitative manner using VAF per mutation per region. Among 14 tumors, 12 tumors had at least one founder mutation from a total of 52 founder mutations, whereas 12 tumors had at least one truncal mutation from a total of 19 truncal mutations. Excluding hypermutators, 52 founder and 19 truncal mutations were identified among all 248 types of mutations found in 12 tumors. In addition, 18 founder mutations (18/52, 34.6%) were also truncal mutations. Importantly, the great majority (18/19, 94.7%) of truncal mutations were founder mutations.

### Intratumor copy number variation

Since a sequencing panel was used to identify single nucleotide variations (SNVs) and insertion-deletion mutations (INDELs), the ability to assess copy number variations (CNVs) was less comprehensive than a whole genome sequence. In fact, using sequencing results from the current cancer panel, the average number and size of detected CNVs was 16.6 events and 19.3 Mb, respectively. We used the ONCOCNV algorithm to characterize large copy number changes from gene panel sequencing (21). CNVs occurring in two arbitrary loci indicated strong correlations (r >0.9) (Fig. 2B, C). In 42 possible combinations from three regions of 14 tumors, the median within patient correlation coefficient was 0.86 (IQR: 0.70-0.92) (Supplementary Fig. S2). Based on the normalized CNV across 42 samples, 184 and 140 genetic regions were identified as having gain and loss of copy number, respectively (Supplementary Fig. S3). Overall, the high correlation among sample regions and notable CNVs suggest that CNVs, including some that have potentially critical functions, likely occurred at a relatively early stage during tumor development.

### Validation of mutations using dPCR

A set of 34 unique mutations for all tumors was selected to monitor tumor burden. dPCR was used to confirm concordance of VAFs between dPCR and NGS. Among the available DNA samples, tumor genetic heterogeneity was evaluable by dPCR in 121/123 (41 mutations × 3 regions) specimens. Among the 121 mutations, 103 had been identified by NGS, whereas the remainder had not. Of the 18 mutations that had not been detected by NGS, 10 (55.6%) were detected by dPCR. Of these mutations, half showed <1% VAF, likely accounting for the discordance. The overall concordance in mutation detection in a binary manner (i.e., presence or absence) between NGS and dPCR was 91.7% (111/121 mutations; Supplementary Data file S1). The VAFs measured by NGS and dPCR showed a good correlation, particularly in the high (>1%) range (r=0.79; Supplementary Fig. S4).

### Proteomic profile of multiregion samples

Two-way unsupervised hierarchical clustering across 42 samples and 293 proteins revealed two major protein clusters with only 6/14 tumors including all multiregion samples from that tumor within the closest cluster (Supplementary Fig. S5A). Although 12/14 tumors had at least one founder mutation, the number or prevalence of the founder mutations did not appear to be closely associated with protein profile. For instance, multi-region samples of the tumor from CC16001, which had eight founder mutations, were separated in the proteomic clusters. In contrast, the CC16016 tumor had two founder mutations consistent with a greater degree of heterogeneity, yet all three samples were located in the same proteomic cluster. These findings suggest that genetic heterogeneity did not dominantly affect protein levels tested in the present study. Notably, levels of Src phosphorylation were high in the CC16033L tumor, whereas levels of Bcl-xL were high in the CC16033R tumor. These two tumors from a single patient suggest that the molecular development of tumors in the same individual may occur via distinct activation pathways that cannot be distinguished based on genetic information alone (Supplementary Fig. S5A). To further investigate the association between gene mutation and levels of the encoded protein, we compared protein levels based on gene mutation status. Among 151 genes and 293 proteins, 43 genes had matched proteins that allowed gene-protein level comparison. All comparisons were carried out using multiregional samples in which specimens were divided into two pieces for gene sequencing and reverse phase protein array (RPPA) (Supplementary Fig. S1). Interestingly, none of the eight pairs demonstrated different levels of proteins in terms of gene mutation status. Although antibodies used in RPPA were not designed against specific mutated proteins, these results suggest that gene mutation does not dominantly contribute to the protein level (Supplementary Fig. S5B).

### ctDNA in preoperative plasma samples

The average amount of preoperative cell-free DNA (cfDNA) in 1 mL plasma was 14.3 ng (range 7.2-34.2). Preoperative tumor specific mutations were detected as ctDNA in 9/12 patients (75.0%) using dPCR. The average VAF of ctDNA for all disease stages assessed by dPCR was 1.02%± 5.1 (±2SD). By Stage, the average VAF of preoperative ctDNA for Stage III and Stage IV A-B was 0.60%±1.32 (±2SD) and 3.67%±10.12 (±2SD), respectively. The detection rates for ctDNA of founder and non-founder mutations were 67.9% (19/28 mutations) and 61.5% (8/13 mutations), respectively; those of truncal mutations and branch mutations were 75.0% (9/12 mutations) and 62.1 % (18/29), respectively (Supplementary Fig. S6, Supplementary Table S5). As expected, the detection rate for ctDNA of founder mutations was higher than that of non-founder mutations, and the detection rates as ctDNA of truncal mutations were higher than those of branch mutations. Among primary tumors with >10% VAF mutations as well as >100 read counts by NGS, 74.0% (77/104) of detected ctDNA were derived from either founder or truncal mutations. Overall, high VAF mutations seemed to be a reasonable surrogate of either founder or truncal mutations.

### ctDNA monitoring by dPCR over time

In contrast to NGS, dPCR offers several advantages, including high sensitivity, rapid turn-around-time, and low cost. These factors allow the frequent monitoring of individual tumor-specific mutations. We propose that ctDNA monitoring as a tumor marker could contribute to: (a) early relapse detection; (b) treatment efficacy evaluation; and (c) non-relapse corroboration. As examples, we consider the following cases.

Patient CC16041 had Stage III rectal cancer and underwent surgery with curative-intent. The tumor from this patient carried a founder mutation for *TP53* (c.524C>T) and a non-founder mutation for *DDR2* (c.442A>T), which were both selected to assess the efficacy of ctDNA monitoring (Fig. 3A). Preoperative ctDNA for both *TP53* and *DDR2* decreased immediately after surgery, with decreases in *TP53* ctDNA levels, which had the highest VAF, being more pronounced than those for *DDR2*. Although levels of both mutated ctDNAs fluctuated during post-operative adjuvant chemotherapy, they both remained low with levels around 0.1% or lower. Strikingly, *TP53* ctDNA showed a subsequent increase after completion of adjuvant chemotherapy. The peak *TP53* ctDNA level occurred on day 409 on which a follow-up CT identified a recurrent lesion at a para-aortic lymph node (Fig. 3A, CT2). Importantly, the increase in *TP53* ctDNA preceded imaging detection of recurrence by 90 days. Subsequent bevacizumab + FOLFILI therapy resulted in a decrease in the size of the recurrent lesion on day 622 as well as a decrease in *TP53* ctDNA (Fig. 3A, CT3). For this patient the concomitant serum tumor marker CEA and DDR2 ctDNA did not reach “positive” levels during the entire treatment course. This suggests that the clone(s) that resulted in the recurrence contained the founder *TP53* mutation in a large fraction, but not the non-founder *DDR2* mutation.

**Figure 3.**
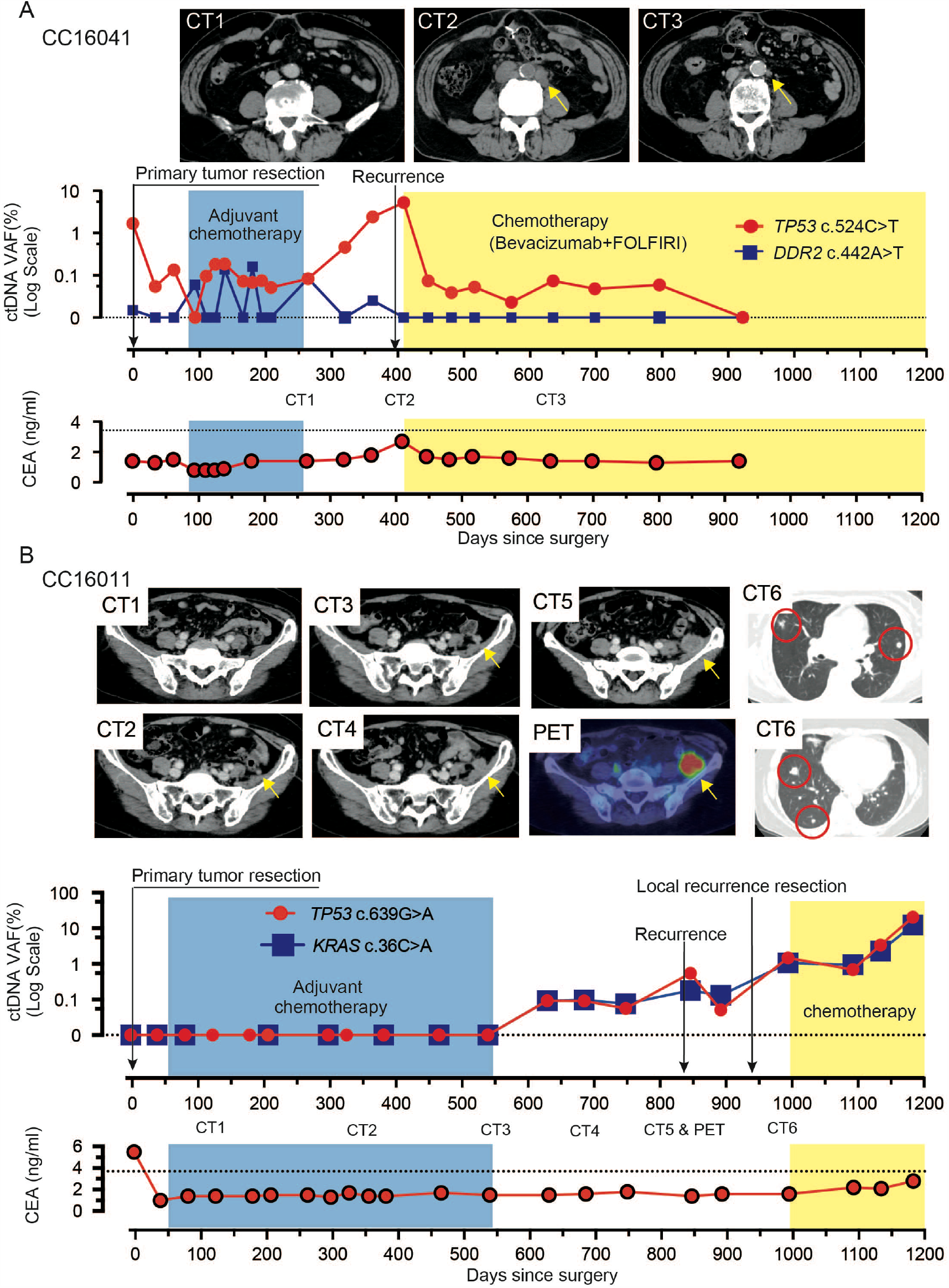
Early relapse prediction by ctDNA monitoring recurrent cases. (A) An aortic lymph node recurrence was noted by CT2 on day 409 after primary tumor resection. As noted in CT3, the size of the aortic lymph node recurrence was reduced on day 622. (B) Levels of ctDNA were undetectable up to day 539 during preoperative and adjuvant chemotherapy. Yellow arrowheads in CT images indicate a suspected local recurrence lesion in the pelvic peritoneum; this diagnosis was confirmed on day 846 (CT5 & PET). Continuous detection of ctDNA was possible for seven months before the recurrence was diagnosed by CT scan. Multiple lung metastases were diagnosed at day 980, after surgery for recurrence in the peritoneum (CT6). The patient began receiving chemotherapy on day 1,015, at which point the ctDNA declined temporarily before re-increasing through day 1,200. PET, positron emission computerized tomography.

Patient CC16011 exhibited a different pattern. ctDNA was not detectable pre-surgery and remained undetectable for >500 days after surgery. ctDNA levels became detectable at day 629 and remained elevated despite surgery to remove a recurrent tumor and subsequent chemotherapy. Importantly, however, the increase in ctDNA preceded to local recurrence detected by CT on day 846, approximately 300 days after adjuvant chemotherapy ended (Fig. 3B). Once again, ctDNA increases also preceded increases in CEA. These observations suggest that individual tumor-specific ctDNA monitoring can provide information on recurrence for patients who underwent tumor resection with curative-intent.

Patient CC16001 demonstrated that ctDNA for three mutations was detectable in preoperative plasma (Fig. 4A). Post-operatively, ctDNA remained undetectable for nearly 1,200 days. CEA levels for this patient dropped to standard levels after primary tumor resection. The estimated 3-year recurrence risk for CC16001 according to the final disease stage was approximately 25% (23). The continuous undetectable levels of ctDNA suggest that this patient was at low risk for recurrence, which was confirmed by a lack of recurrence for 1000 days. In the present case series, six additional patients (CC16008, CC16016, CC16023, CC16034, CC16042, and CC16043) with >Stage IIIA had a similar pattern to patient CC16001, whereby levels of ctDNA that were detectable pre-therapy immediately decreased after tumor resection and remained undetectable for approximately 1,000 days (Supplementary Fig. S7).

**Figure 4.**
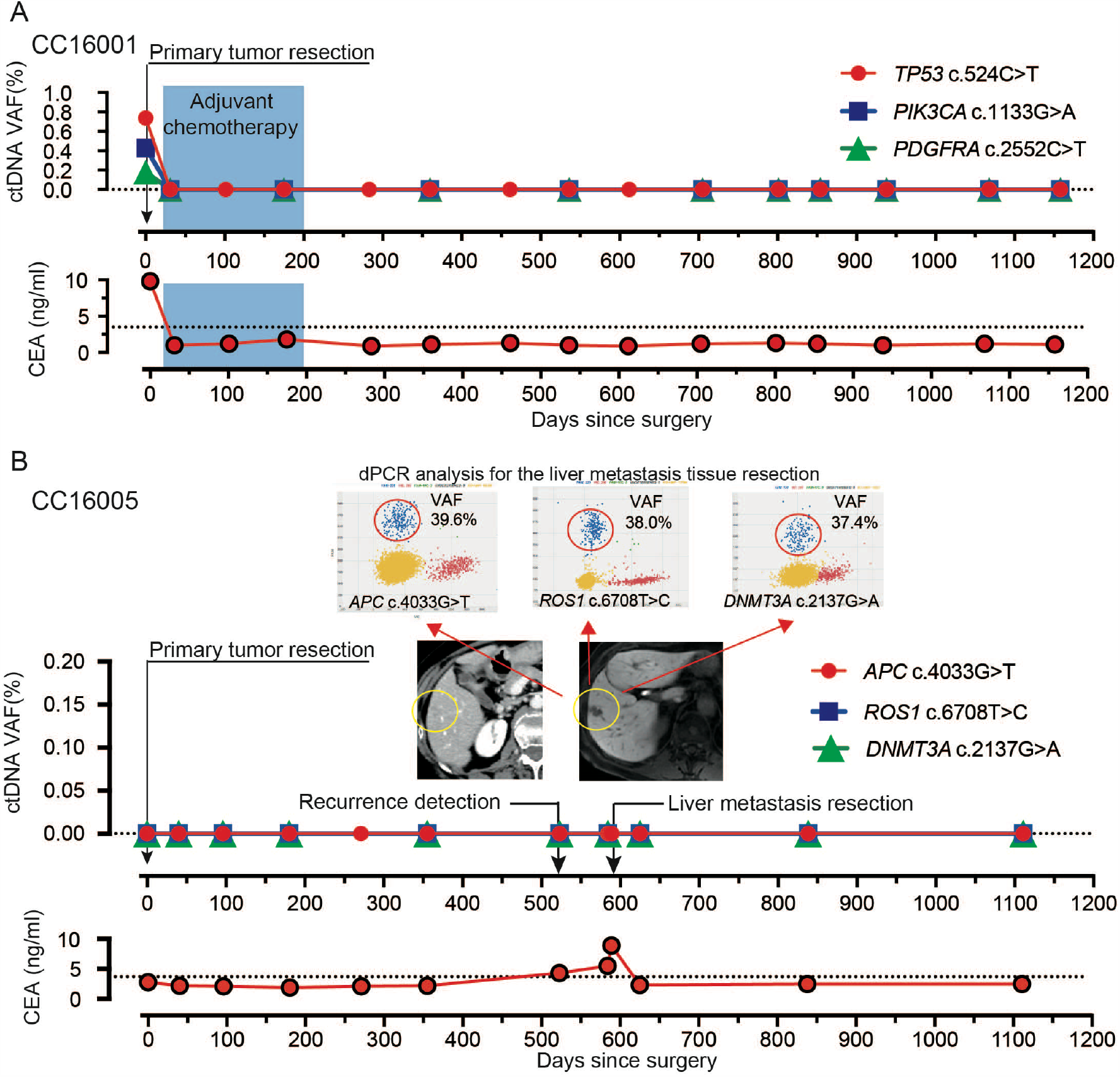
Undetected ctDNA monitoring by dPCR. (A) A case with no recurrence after primary colorectal cancer resection. (B) A patient was diagnosed with a single liver metastasis 1.5 years after the first operation. Three mutations in ctDNA were not detectable pre-operatively, but were detected by dPCR in resected metastatic tissue.

Patient CC160005 demonstrated another pattern. Despite concordance of mutations detected in the primary tumor and in a recurrent metastatic liver lesion, ctDNA was not detected throughout the patient’s course of management (Fig. 4B). Patient CC16035 had also undetectable ctDNA levels at pretreatment and throughout the treatment period with three mutations (Supplementary Fig. S8). While CC16035 did not recur, patient CC16005 developed a metastatic lesion that was not accompanied by an increase in ctDNA levels.

Finally, patient CC16033 demonstrated another interesting pattern. CC16033 had synchronous double right (i.e., cecum) and left (i.e. sigmoid) colon tumors concomitant with metastases to the lung and liver (Fig. 5A). Local resection of both primary tumors was performed, followed by chemotherapy. Metastatic lesions of the lung and the liver showed marginal responses to the two lines of chemotherapy (Fig. 5B). Interestingly, both tumors had founder mutations of the same gene set (i.e., *TP53, APC*, and *KRAS*) but the mutation positions were all different. Following primary tumor resection, ctDNA levels of one set of founder mutations identified from the left colon tumor decreased and remained undetectable (Fig. 5C). However, ctDNA levels of another set of three founder mutations from the other (i.e., right) lesion did not decrease after surgery, but did decrease during chemotherapy to near the detection limit (0.1%) of VAF. The ctDNA levels of the three founder mutations from the right tumor remained low until approximately 600 days post-surgery when they began to increase and remained high until death. Importantly, the dynamics of two sets of three founder mutation levels remained concordant throughout the course of therapy, suggesting that they were from the same clone.

**Figure 5.**
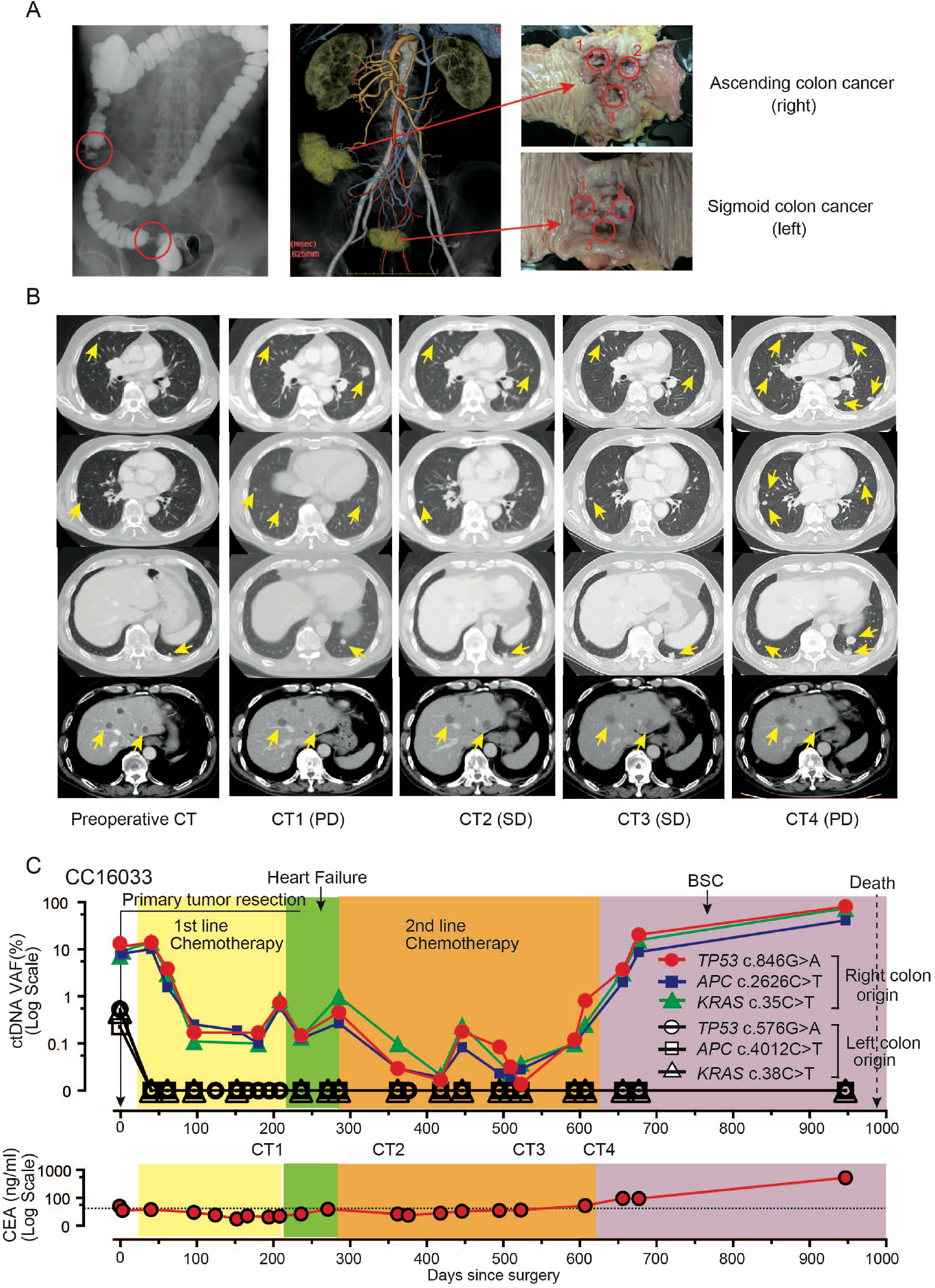
Longitudinal ctDNA monitoring in patients with multiple cancer. (A) A macroscopic abdominal view of a patient having two tumors in the cecum (right) and sigmoid colon (left). Multi-regional samples were taken from each tumor. (B) CT imaging studies of the pre- and post-operative course for synchronous metastases to liver and lung. Yellow arrows indicate metastatic lesions. (C) A set of three mutations identified from the right colon tumor were detected as ctDNA after resection of both primary tumors and reflected the effect of chemotherapy. Another set of three mutations identified from the left colon tumor was only detected in pre-operative plasma as ctDNA and subsequently remained undetectable. PD, progressive disease. SD, stable disease. BSC, best supportive care.

Our present series allowed us to estimate the clinical validity of ctDNA (24) in terms of the abovementioned categories (i.e., early relapse prediction, treatment efficacy evaluation, and non-relapse corroboration). Overall, for 10 of the 12 patients analyzed, there was information available in the personalized ctDNA analysis that could result in patient benefit with a good contrast to conventional serum tumor marker, CEA (Fig. 6).

**Figure 6.**
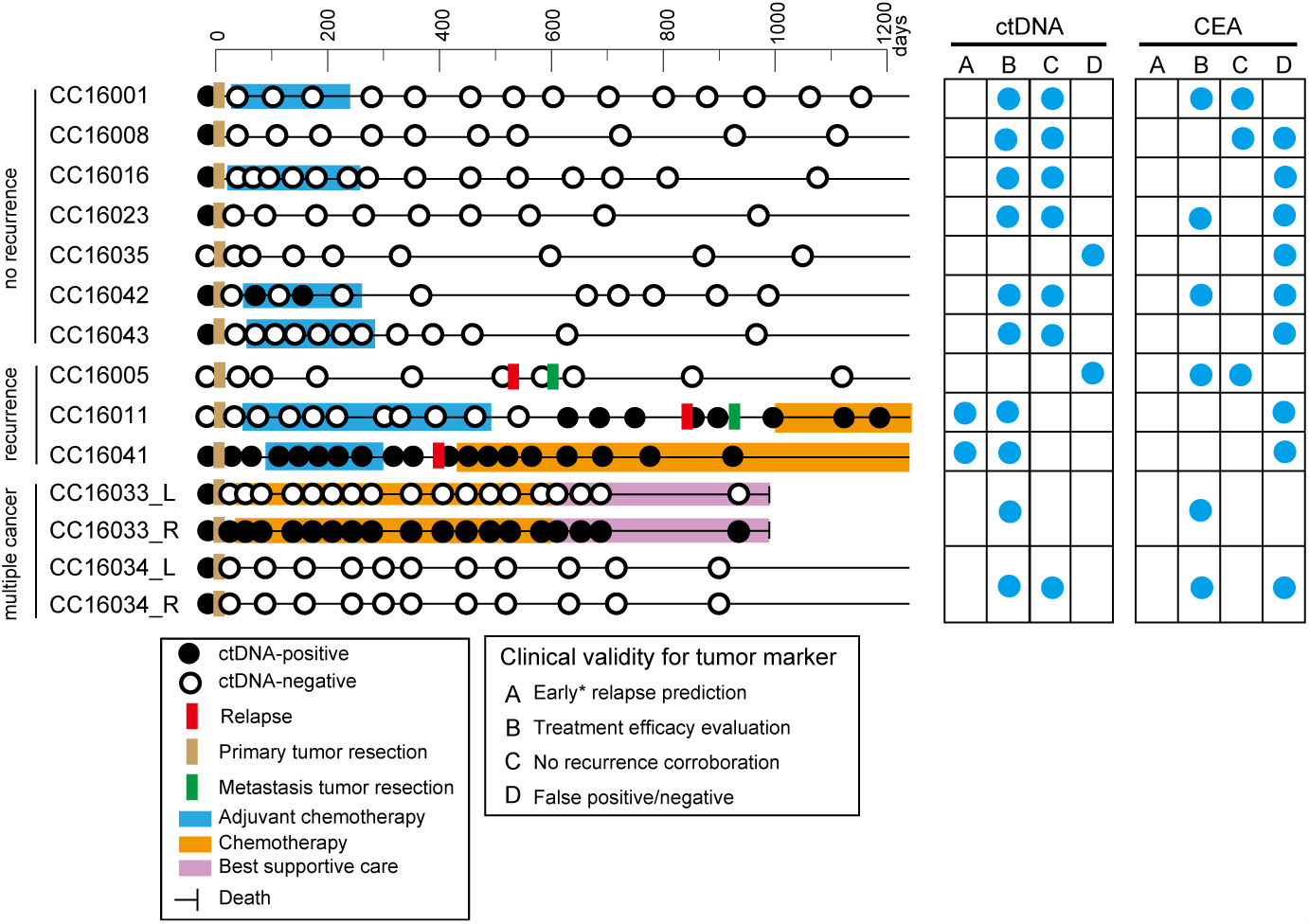
Longitudinal ctDNA status and individual clinical validities. ctDNA status and clinical information are illustrated on the horizontal lines by case. Blue circles in the right grid indicate representative clinical validities of ctDNA and CEA. (A) early relapse prediction; (B) treatment efficacy evaluation; (C) non-relapse corroboration; and (D) false positive/negative.

## Discussion

The ultimate clinical utility of tumor markers is their ability to provide unique information that can alter patient management leading to improved survival (24). Therefore, subjects who are most likely to benefit from tumor marker data are those with an intention-to-treat approach designed to reduce tumor burden and increase survival rate (25). In a search for clinically useful tumor markers for patients with advanced-stage CRC, we examined ctDNA in terms of tumor heterogeneity, proteogenomics, and detection limit. Unique, practical information can be obtained from ctDNA including: (a) early relapse detection, (b) treatment efficacy evaluation; and (c) non-relapse corroboration. Our results suggest that the presence of high VAF mutations in a tumor can be a practical surrogate for founder mutations that are likely to be detected as ctDNA(26).

To establish ctDNA as a clinically useful marker, further biological investigations are needed. In contrast to studies that focused on tumor phylogeny, here we assessed the genetic heterogeneity of tumors using a simple, minimum number (i.e., three) multiregion sequencing, which was chosen to represent a practical number for standard clinical practice. The objective of the multi-region sequencing was to identify founder and truncal mutations. While a “founder” mutation is defined in a binary manner (i.e., presence or absence), “truncal” mutations take VAF (i.e., continuous variables) into account. We demonstrated that dPCR was able to detect the presence of mutations that had not been identified by NGS, which resulted in more founder mutations than expected. This observation may suggest, under the binary distinction, that if genetic heterogeneity is assessed with techniques whose quantitative detection limit is less than 0.1%, then the prevalence of the genetic heterogeneity may be less than previous studies exhibiting a lower depth of analysis have suggested (8,27,28). Therefore, it is reasonable to use either founder or truncal mutations at the NGS sensitivity level for ctDNA monitoring, since these mutations should have a higher chance to be released from anywhere within the tumor. However, it is impractical to run multi-region sequencing in every single patient. In practice, the present results suggest that a high VAF may be a good surrogate of this class of mutation (26). In fact, using high VAF mutations from primary tumors for ctDNA monitoring, we showed that ctDNA provided a 3-6 month lead time compared to conventional imaging examinations for early relapse prediction. Moreover, in daily practice, both treatment efficacy and non-relapse corroboration could be assessed with greater frequency, which is likely to extend the lead time in case treatment action is needed.

The potential role of ctDNA as a basis for selection of molecular targeting drugs remains unclear beyond a subset of therapies. Currently, some established mutation-drug pairs have shown utility, such as a tyrosine kinase inhibitor (TKI) (29) for NSCLC, immune checkpoint antibodies (30), B-Raf inhibitors for melanoma (31,32), and an anti-human epidermal growth factor receptor type (HER2) antibody for breast and gastric cancer (33,34). Although some matched therapies are associated with improved survival (35), the majority of the therapeutic targets are proteins and there is limited information about whether gene mutations are a direct cause of functional deficiency of the encoded protein. We were able to assess 42 gene-protein pairs using sequencing and RPPA. Overall, there was no clear association between gene mutational status and levels of the corresponding protein.

There are several limitations to the present study: (i) the number of enrolled patients (n=12) was relatively small; (ii) the number of regions sampled (three per tumor) may not have allowed estimation of tumor-wide clonal heterogeneity; (iii) the number of mutations monitored that were specific to each patient was limited and was not designed to assess clonal changes; and (iv) the survey period (median 965 days) may not be sufficiently long to evaluate late recurrence. Nevertheless, the ctDNA results provide clues to pursue further areas of study with a focus on addressing the current limitations.

In summary, dPCR allows frequent and rapid assays with at least a 10-fold lower detection limit compared to NGS (6). A limited set of personalized mutation targets was sufficient to monitor tumor burden. Tumor genetic heterogeneity does not appear to represent a major obstacle for ctDNA monitoring if an appropriate high VAF somatic mutation is selected. Further interventional prospective studies are needed to confirm that ctDNA monitoring provides an effective approach for extending patient survival.

## Data Availability

Currently in the process of data uploading to DDBJ.

## Acknowledgments

We thank all the patients who participated in this study. We also thank K. Takashimizu, H. Fujii, T. Matsuo, M. Ikeda, and Y. Ohmori for expert technical assistance.

## Author contributions

M.Y., F.E., T.I., S.S.N. designed the study; M.Y., K.S., T.K., K.O. performed colorectal cancer treatments, diagnosed clinical recurrence, and made treatment evaluations; R.S., T.S., L.L. assessed the primary tumor tissue; M.F., Z.J., T.H., H.N. performed NGS and CNV analysis and constructed a phylogenic tree for primary tumors; D.S., Y.L., G.M., S.S.N. performed RPPA analysis; M.Y., T.I., N.S., F.E. performed genomic DNA extraction and dPCR analysis; M.Y., G.B.M., S.S.N. contributed to writing of the manuscript.

